# Fast, accurate and robust sparse-view CT reconstruction via residual-guided Golub-Kahan iterative reconstruction technique (RGIRT)

**DOI:** 10.1101/2023.02.24.23286409

**Authors:** Jianru Zhang, Zhe Wang, Tuoyu Cao, Guohua Cao, Wuwei Ren, Jiahua Jiang

## Abstract

Reduction of projection views in X-ray computed tomography (CT) can protect patients from over exposure to ionizing radiation, thus is highly attractive for clinical applications. However, image reconstruction for sparse-view CT which aims to produce decent images from few projection views remains a challenge. To address this, we propose a Residual-guided Golub-Kahan Iterative Reconstruction Technique (RGIRT). RGIRT utilizes an inner-outer dual iteration framework, with a flexible least square QR (FLSQR) algorithm implemented in the inner iteration and a restarted iterative scheme applied in the outer iteration. The inner FLSQR employs a flexible Golub-Kahan (FGK) bidiagonalization method to reduce the dimension of the inverse problem, and a weighted generalized cross-validation (WGCV) method to adaptively estimate the regularization hyper-parameter. The inner iteration efficiently yields the intermediate reconstruction result, while the outer iteration minimizes the residual and refines the solution by using the result obtained from the inner iteration. Reconstruction performance of RGIRT is evaluated and compared to other reference methods (FBPConvNet, SART-TV, and FLSQR) using realistic mouse cardiac micro-CT data. Experiment results demonstrate RGIRT’s merits for sparse-view CT reconstruction in high accuracy, efficient computation, and stable convergence.

## 1. Introduction

X-ray computed tomography (CT) has become an indispensable tool in various fields, including industrial inspection, security checks, and medical diagnosis of a wide range of diseases [1]. However, traditional CT scanning requires a dataset from a large number of projection angels, which can expose patients to excessive ionizing radiation, increasing the risk of cancer [2]. To lower the CT radiation dose in clinical practice, sparse-view CT scanning with a small number of projection views was proposed. Sparse-view CT has the significant advantage of reducing the radiation dose proportionally to the number of projection views.

Sparse-view CT could also improve scanning speed and hence temporal resolution by shortening data acquisition time, making it particularly useful for some dynamic imaging tasks, such as cardiac micro-CT imaging for small animals [3].

Despite the potential benefits of sparse-view CT, one significant challenge is image reconstruction due to insufficient projection data. Reconstructing images from under-sampled projection data is an ill-posed inverse problem [4]. Traditional methods such as filtered back projection (FBP) are not applicable, as they produce highly degraded images with severe streaking artifacts [5]. Model-based iterative reconstruction methods have been quite successful in addressing the problem of insufficient projection views in sparse-view CT. Some classical iterative algorithms include gradient based methods and the Kaczmarz family of algorithms, such as algebraic reconstruction technique (ART) [6, 7], simultaneous iterative reconstruction technique (SIRT) [8], and simultaneous algebraic reconstruction technique (SART) [9]. Despite being a significant improvement over traditional methods, these iterative reconstruction algorithms are still limited by their low reconstruction accuracy and slow convergence speed.

In addition to the iterative methods mentioned above, there has been a growing interest in using deep learning (DL)-based methods for sparse-view CT reconstruction, as they are capable of leveraging large amounts of CT data [10]. DL methods can be trained for the sparse-view CT image reconstruction task in the sensor domain [11], image domain [12], or dual domains [13]. After the training, the trained DL model can then be used to reconstruct sparse-view projection data either with or without the assistance of traditional or iterative methods. However, the training process for DL models can be computationally expensive, and the resulting network models usually lack explainability and generalizability. To address these limitations, there is a current trend toward designing hybrid DL models that combine the strengths of deep learning and model-based iterative reconstruction [14-16]. Therefore, it is beneficial to research for model-based iterative reconstruction methods that can generate accurate reconstruction results quickly and stably.

In iterative reconstruction of sparse-view CT, it is crucial to introduce regularization with some prior knowledge. By introducing reasonable priors with more accurate modeling of the imaging system and measurement data, reconstruction quality can be improved. One effective regularization method is total variation (TV) regularization, which can suppress noise and preserve sharp edges by minimizing the overall statistical variation of the target image [17]. Multiple variants of TV regularization have been proposed [18-20]. However, TV regularization can result in patchy artifacts due to its assumption that the image is piecewise constant. Additionally, compressed sensing theory [21, 22] has led to the development of many sparse regularization-based iterative reconstruction algorithms, such as *ℓ*_1_ norm or generalized *ℓ*_*p*_(0 ≤ *p* ≤ 1) norm regularization. These methods improve image quality by reducing artifacts and preserving high-frequency information. However, optimization problems with *ℓ*_1_ norm penalty can be computationally challenging due to difficulties in estimating the regularization parameter and non-differentiability at the origin, requiring expensive nonlinear or iterative reweighted optimization schemes.

To quickly and stably solve the *ℓ*_1_ regularization problem in sparse-view CT reconstruction, a variety of algorithms and acceleration techniques have been developed. One classical algorithm is the fast iterative shrinkage-thresholding algorithm (FISTA), which achieves a faster global convergence rate by adopting the first order gradient algorithm while retaining the simplicity of the traditional iterative shrinkage-thresholding algorithm (ISTA) [23]. Other similar works based on the FISTA principle have also been reported [24, 25]. Additionally, dictionary learning, which assumes that the signal can be decomposed into a sparse representation in a redundant basis of patterns (also known as sparse coding), is also an effective approach to overcome the non-differentiability problem and reduce dimension of the original inverse problem [26-28]. Although these methods can improve the reconstruction accuracy to some extent, they still face several challenges, such as high memory cost and long computation time, and difficulty in selecting an appropriate regularization parameter.

To overcome these challenges, flexible least square QR (flexible LSQR or FLSQR) has been recently proposed to solve the regularization problem in iterative reconstruction for sparse-view CT. FLSQR has shown promising results in sparse-view CT reconstruction of Shepp-Logan numerical phantom [29]. This method is a variation of the basic Krylov method, which works by expanding an approximation subspace for the solution and solving a projected least squares problem at each iteration [30]. FLSQR offers two key advantages. Firstly, it introduces an efficient projection method, called flexible Golub-Kahan (FGK) process [31], to reduce the size of the original large-scale regularized problem, making it computationally efficient. Secondly, the regularization parameter in FLSQR can be selected automatically, efficiently, and adaptively in an iterative manner, enabling stable convergence. Despite the success of FLSQR in numerical simulations, its practical applications in realistic CT reconstruction have not been fully explored.

Building on the success of FLSQR, we propose a new iterative reconstruction technique for sparse-view CT called Residual-Guided Golub-Kahan Iterative Reconstruction Technique (RGIRT), which retains the speed advantage of FLSQR while achieving higher reconstruction accuracy. RGIRT utilizes an inner-outer iteration framework, where FLSQR is implemented in the inner iteration and a restarted iterative scheme is applied in the outer iteration. The outer iteration is responsible for minimizing the residual and ensuring convergence, which improves reconstruction accuracy. In the inner iteration, we repeatedly solve the projected problem with the solution space dimension increasing from 1 to the maximum iteration number for the inner loop when the inner solver reaches its stopping criteria. However, this dimension is still significantly smaller than the ultimate projected problem dimension in FLSQR. Therefore, this inner-outer iteration scheme not only enhances reconstruction accuracy but also reduces memory cost and accelerates the reconstruction process.

The remainder of the paper is organized as follows. Section 2 mainly introduces the derivations and specific operations of our proposed RGIRT algorithm and the data acquisition details of the mouse cardiac micro-CT experiment, which provides the realistic raw data for reconstruction. In section 3, reconstruction results from the mouse micro-CT data are presented, and the effectiveness of our proposed method is evaluated both qualitatively and quantitatively. Finally, discussion and conclusion are presented in section 4.

## 2. Method and Experiments

### 2.1 Residual-guided Golub-Kahan iterative reconstruction technique

#### 2.1.1 Inner-outer iterative scheme

The ill-posed nature of sparse-view CT image reconstruction calls for the use of regularization to stabilize the inverse process. The mathematical model can be formulated as

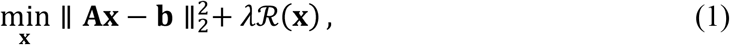

where **x** ∈ *R*^*N*^ is the discrete image array; **b** ∈ *R*^M^ is the corresponding measurement data (sinogram) that carries noise; **A** ∈ *R*^M×*N*^ is the system matrix that models the forward process; M is the number of source-detector pairs, *N* is the number of CT image grids. *ℛ*(**x**) denotes the regularization term, and *λ* (*λ* > 0) is the regularization parameter that balances the data fidelity term 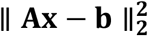 and the regularization term *ℛ*(**x**). The primary framework of our method is based on inner-outer iterations (as shown in Fig. 1), where FLSQR is performed in the inner iterations to solve a subproblem until convergence, and the outer iteration refines the intermediate solution to the main problem (1) heuristically using the results from the inner iteration. Given an initial guess **x**^(0)^, we define the inner problem at the (*ℓ* + 1)-th outer iteration as

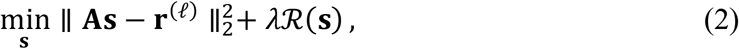

where **r**^(*ℓ*)^ = **b** − **Ax**^(*ℓ*)^ represents the residual at the *ℓ*-th outer iteration and **x**^(*ℓ*)^ is the approximated solution at the *ℓ*-th outer iterative; **s** represents the correction that will be used for updating **x**^(*ℓ*)^ in the outer iteration. Subsequently, we obtain the (*ℓ* + 1)-th outer iterative result **x**^(*ℓ*+1)^ = **x**^(*ℓ*)^ + **s**. Although there are multiple choices for *ℛ*(**s**), in this work we use *ℓ*_1_ regularization, which is defined as *ℛ*(**s**) = ∥ **s** ∥_1_. Furthermore, we proved the convergence of our proposed algorithm. We show that incorporating the correction in the outer iteration facilitates a gradual reduction of the residual, thereby mitigating the concern regarding semi-convergence. This can be expressed as

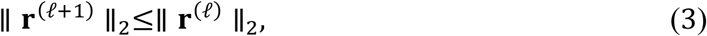

where the proof is provided in Appendix A.

**Fig. 1.**
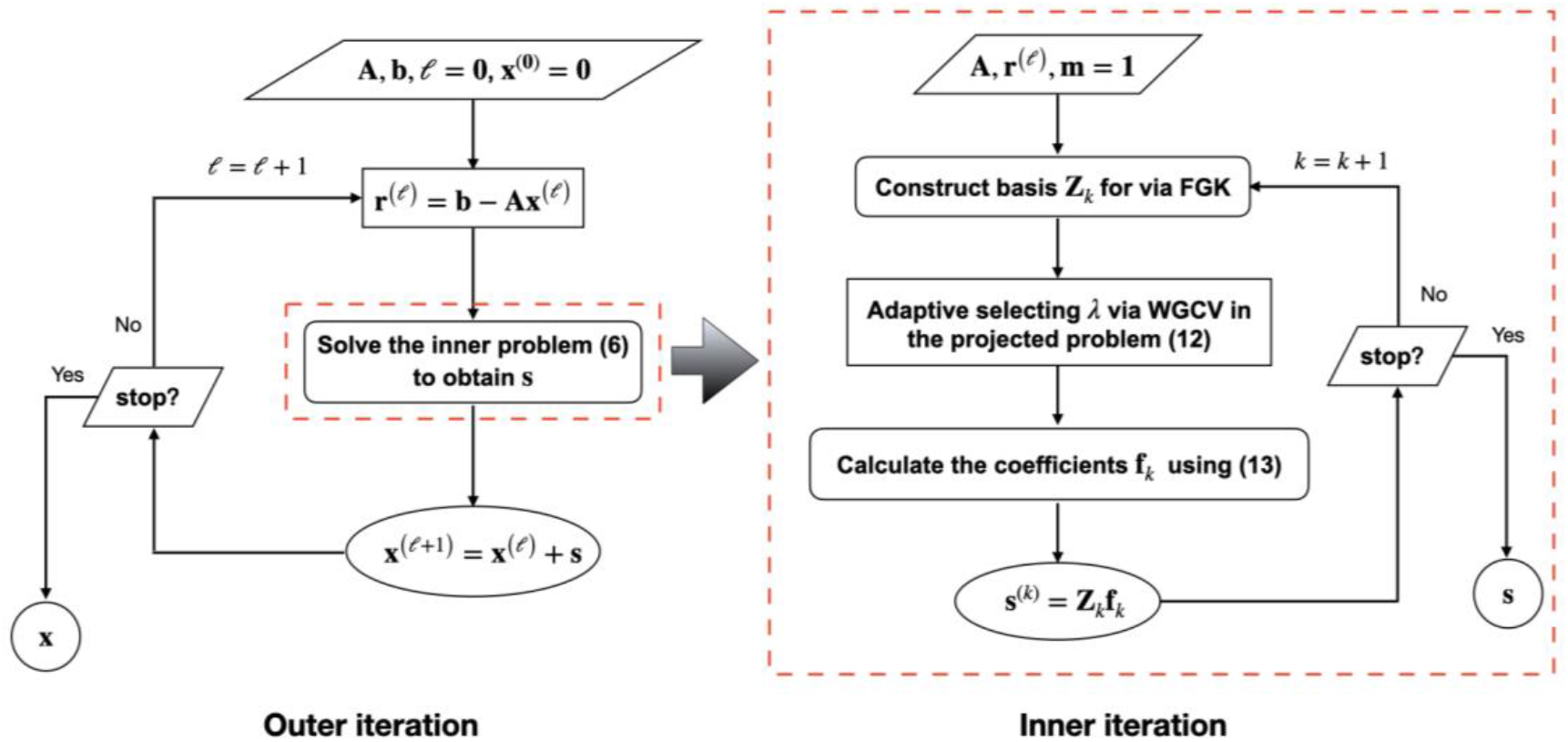
The framework of RGIRT. RGIRT minimizes the residual in the outer iteration and employs the FGK process in the inner iteration to reduce its dimension adaptively.

#### 2.1.2 Optimization via iterative reweighted norms

Iterative reweighted norms (IRN) method has proven effective in solving inverse problems with *ℓ*_1_ regularization constraint, by approximating *ℓ*_1_ regularization with iteratively updated weight matrix [32, 33]. The first step of IRN approach is to reduce *ℓ*_1_-regularized problem to a sequence of least-squares problems involving a weighted *ℓ*_2_ norm.

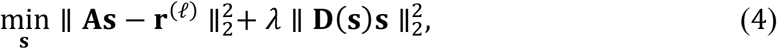

where

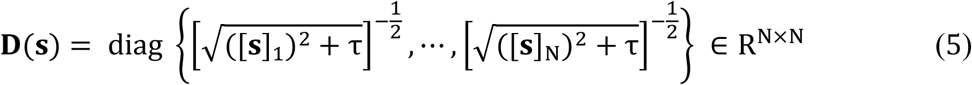

with [**s**]_i_ denoting the *i*-th entry of **s** and a small threshold *τ* > 0. To avoid nonlinearities of solving (4), we follow the common practice of approximating **D**(**s**) by the weighted matrix at the *k*-th inner iteration **D**_***k***_ = **D**(**s**^(*k*−1)^), where **s**^(*k*−1)^ is an approximation of the solution at the (*k* − 1)-th inner iteration. Notice that seeking **s**^(*k*)^ in (4) requires solving a large optimization problem with *N* unknowns at each iteration. To ameliorate the computational burden, we adopt the FLSQR method that can reduce problem dimension and automatically estimate regularization parameter.

#### 2.1.3 Solution for the inner problem via FLSQR

The essence of FLSQR lies in an iterative projection scheme, encompassing two main stages in each iteration. First, we generate a basis (a set of vectors) for the solution by exploiting the FGK process [29]. Second, we compute the coefficients of the basis by solving an optimization problem in the projected subspace, where the regularization parameter can be estimated automatically via weighted generalized cross validation (WGCV) [34].

During the implementation of FGK process in the first stage, at the *k*th iteration we obtain

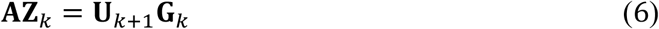

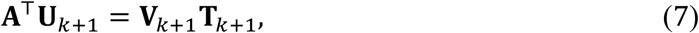

where **G**_k_ ∈ R^(k+1)×k^ is upper Hessenberg and **T**_k+1_ ∈ R^(k+1)×(k+1)^is upper triangular; **Z**_k_ = [**D**_1_^−1^***v***_1_, **…**, **D**_***k***_^−1^***v***_k_] ∈ R^*N*×k^ whose column vectors are the basis for the solution of **s**^(*k*)^; ***V***_*k*+1_ = [***v***_1_ **… *v***_*k*+1_] ∈ *R*^*N*×(*k*+1)^ and ***U***_*k*+1_ = [***u***_1_ **… *u***_*k*+1_] ∈ *R*^*M*×(*k*+1)^ satisfy the orthogonality condition (in exact arithmetic)

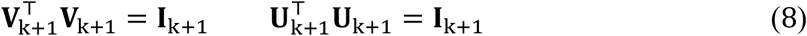

with identity matrix **I**_k+1_ ∈ R^(k+1)×(k+1)^. Then, we can convert problem (4) to

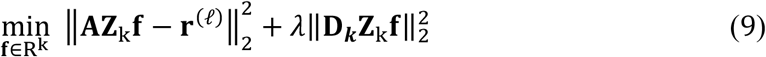

and approximate the solution of (4) via the relations **s**^(*k*)^ = **Z**_*k*_**f**_*k*_ where the basis coefficient **f**_*k*_ ∈ R^k^ is the solution to (9). To further simplify the problem (9), we perform QR factorization of **D**_***k***_**Z**_k_ = **Q**_Z,k_**R**_Z,k_ [35], where **Q**_Z,k_ has orthonormal columns and **R**_Z,k_ is an upper triangular matrix. Plugging the factorization results from equations (6) and (8) into problem (9), we arrive at a small-sized optimization problem in the projected subspace at the *k*-th iteration

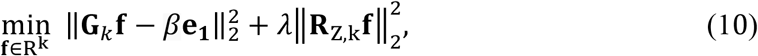

where *β* = ∥ **r**^(*ℓ*)^ ∥_2_ is a scalar, **e**_1_ = [1,0, **…**, 0]^⊤^ ∈ R^k^. Note that the size of the projected problem (10) is (*k* + 1) by *k*. Therefore, RGIRT alleviates the computational complexity by transforming the challenge of solving *N* unknowns of **s** in the full-sized problem (4) into that of solving *k* unknowns of **f**_*k*_ in the small-sized projected problem (10). After applying WGCV in (10) to estimate the regularization parameters *λ*, we can determine the optimal coefficient

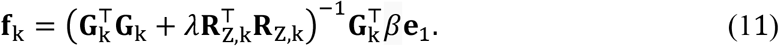

Finally, when the stop criterion of the inner iteration is satisfied, we obtain the solution of problem (1) at the (*ℓ* + 1)-th iteration

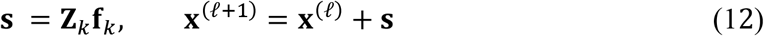

A general overview of RGIRT is illustrated in Fig. 1. Meanwhile, the implementation details of RGIRT are provided in Algorithm 1, where *δ*_*tol*_ is the tolerance given by the user, and ζ_in_, ζ_out_ are the maximum number of the inner and outer iterations, respectively.

##### Algorithm 1

**R**esidual-guided Golub-Kahan Iterative Reconstruction Technique (RGIRT)

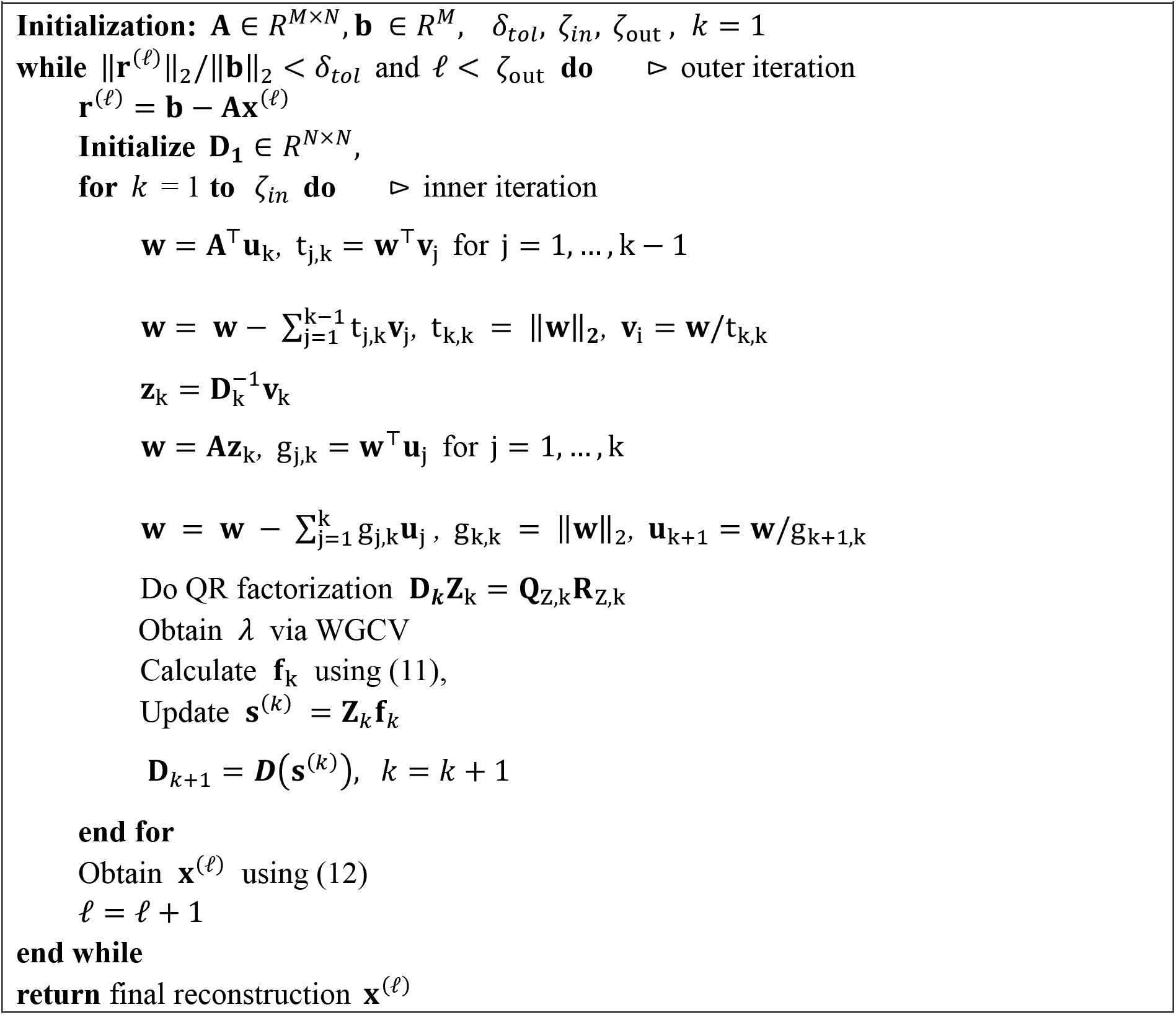

### 2.2 Mouse cardiac Micro-CT data

The sinogram data used in this study were experimentally collected from a mouse cardiac scan using a customized micro-CT scanner [36]. The scanning parameters were 50 kVp anode voltage, 2 mA anode current, 1024 detector elements of 0.76mm pixel pitch for each detector row. The source-to-detector distance and source-to-object distance are 185.03 mm and 141.52 mm, respectively. A total of 400 projections were acquired with a step angle of 0.5°, resulting in a total scan angle of 200° for a short-scan mode in the fan-beam geometry. The 400 projection views were acquired in the step-and-shoot mode with prospective gating to both the respiratory and cardiac signals of the mouse subject under free-breathing condition, so that all the projection views were acquired at the same phase of the cardiac and respiratory cycles. The full 400 projection views were reconstructed by FBP to yield the reference ground-truth image. Then, the sparse projection views, that is, the 100 (or 75, 57, and 39) projection views evenly selected out of the full 400 views were reconstructed by different reconstruction methods to produce the corresponding images for evaluation.

### 2.3 Performance evaluation

We chose four different methods for comparisons including FBP, FBPConvNet [37], SART-TV, and FLSQR [29]. FBP relies on the Radon transform and its inverse, usually viewed as much faster than iterative methods. FBPConvNet is a post-processing DL model based on convolutional neural network (CNN). The SART-TV and FLSQR are both iterative methods. In particular, FLSQR is the same algorithm as our inner iteration but without the outer restarted iterative scheme. For SART-TV, we used a value of 2.5 × 10^−3^ for the regularization parameter. For FBPConvNet, the training dataset consists of 300 mouse cardiac micro-CT images, the validation dataset consists of 100 images, and the testing image is same one as the image reconstructed by other methods. Meanwhile, we used mean square error (MSE) as the loss function and employed the Adam [38] optimizer and a learning rate of 1 × 10^−4^ during network training. The MSE loss function can be written as

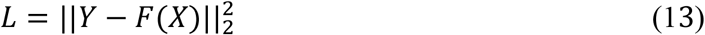

where Y is the ground-truth reference images reconstructed from the full 400 projection views by using FBP, X is the images reconstructed from the sparse-view sinograms via FBP, and F(X) stands for the images post-processed by FBPConvNet.

As for quantitative measures, we used MSE, structural similarity index measure (SSIM) [39], peak signal-to-noise ratio (PSNR), and computational time to evaluate the performance of each algorithm. To evaluate the reconstruction performance of different algorithms under various sparse-view-sampling conditions, CT images were reconstructed from a range of different numbers of views (100, 75, 57, and 39 views, respectively).

## 3. Results

### 3.1 Mouse cardiac micro-CT images reconstructed by different methods

The reconstructed images under various view-sampling conditions of the cardiac micro-CT are shown in Fig. 2. We can observe severe streaking artifacts appears in the images obtained from the FBP algorithm. For the FBPConvNet and SART-TV methods, we tuned regularization parameters at full stretch to balance the effect of artifact removal and image resolution follow the examples from the original papers. We can see that the FBPConvNet, SART-TV, and FLSQR methods can suppress the streaking artifacts to different extents, but their reconstructed images exhibit some over-smoothing effects and the image details were somewhat blurred. Through visual inspection, our RGIRT method provides the best visual image quality in artifact removal and preserving small anatomical details.

**Fig. 2.**
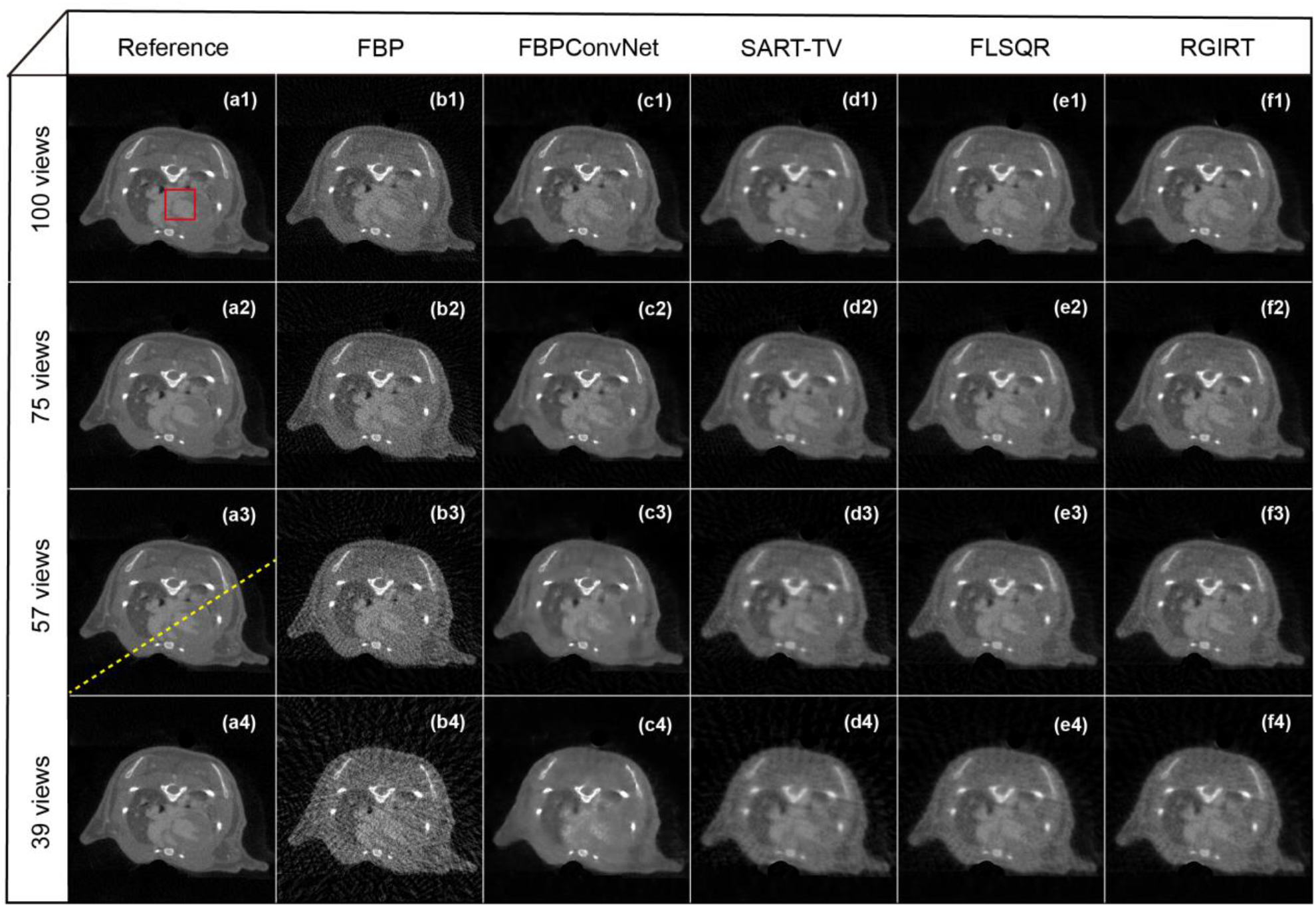
Reconstruction results for a mouse cardiac micro-CT dataset using different methods. Columns from left to right are the full-view reference image and sparse-view image reconstructed via FBP, FBPConvNet, SART-TV, FLSQR and RGIRT, respectively. Images from top to bottom show reconstruction results from 100, 75, 57, and 39 projection views, respectively.

More detailed visual comparison is shown in Fig. 3, where the red box region of Fig. 2 (a1) is enlarged to examine the thin interventricular septum more closely. The interventricular septum (indicated by the blue arrow in the reference image of Fig. 3) is not discernable at all from the sparse-view CT image reconstructed by FBP. For the images reconstructed by FBPConvNet, it is somewhat visible from the 100-view case but barely visible from the remaining fewer-view (e.g. 75, 57 and 39 views) scenarios. The same interventricular septum is visible from the images reconstructed from SART-TV, FLSQR, and RGIRT, but a close look reveals that the interventricular septum reconstructed by SART-TV and FLSQR are a little bit blurry due to the over-smoothing from those two methods. The interventricular septum reconstructed by our RGIRT method seems most clear, especially when examining the small structure details. Moreover, the contrast between the interventricular septum and the neighboring iodine-filled ventricles appears clearer than that from the other methods, which indicates RGIRT is capable of preserving soft tissue contrast and suppressing noise. Judging from Fig. 3, as the sparsity changes from 100 views to 75, 57 and 39 views, the superiority of our RGIRT method becomes even more obvious.

**Fig. 3.**
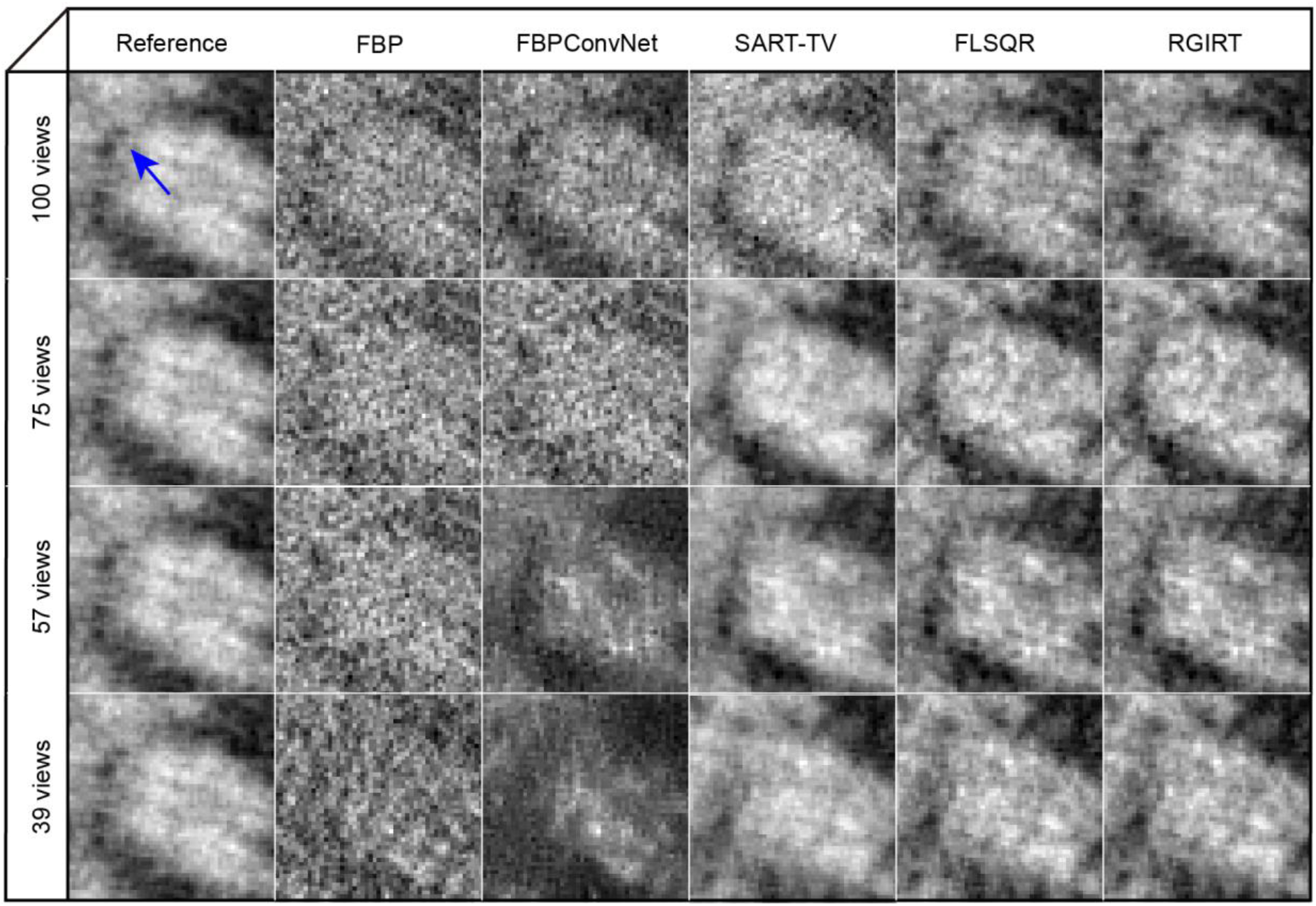
The zoomed region marked by the red box in Fig. 2(a1). Columns from left to right are zoomed images from the reference image and sparse-view images reconstructed via FBP, FBPConvNet, SART-TV, FLSQR and RGIRT, respectively. Images from top to bottom show zoomed region for 100, 75, 57, and 39 projection views, respectively. Blue arrow indicates a thin interventricular septum.

The effectiveness of RGIRT is further demonstrated in the difference images shown in Fig. 4, which were calculated as the absolute difference between the reference image and the sparse-view images in Fig. 2. In these difference images, the darker the color, the larger the error. We can see that the difference image of RGIRT has the smallest overall difference, which confirms RGIRT’s superiority in preserving the image details. Interestingly, comparing the difference images between FBPConvNet and RGIRT, we found that RGIRT is better at preserving the soft tissues, while FBPConvNet is better at preserving the bone structures. Because the heart is a soft-tissue organ without bone structure, our RGIRT method is indeed better suited for sparse-view cardiac CT reconstruction.

**Fig. 4.**
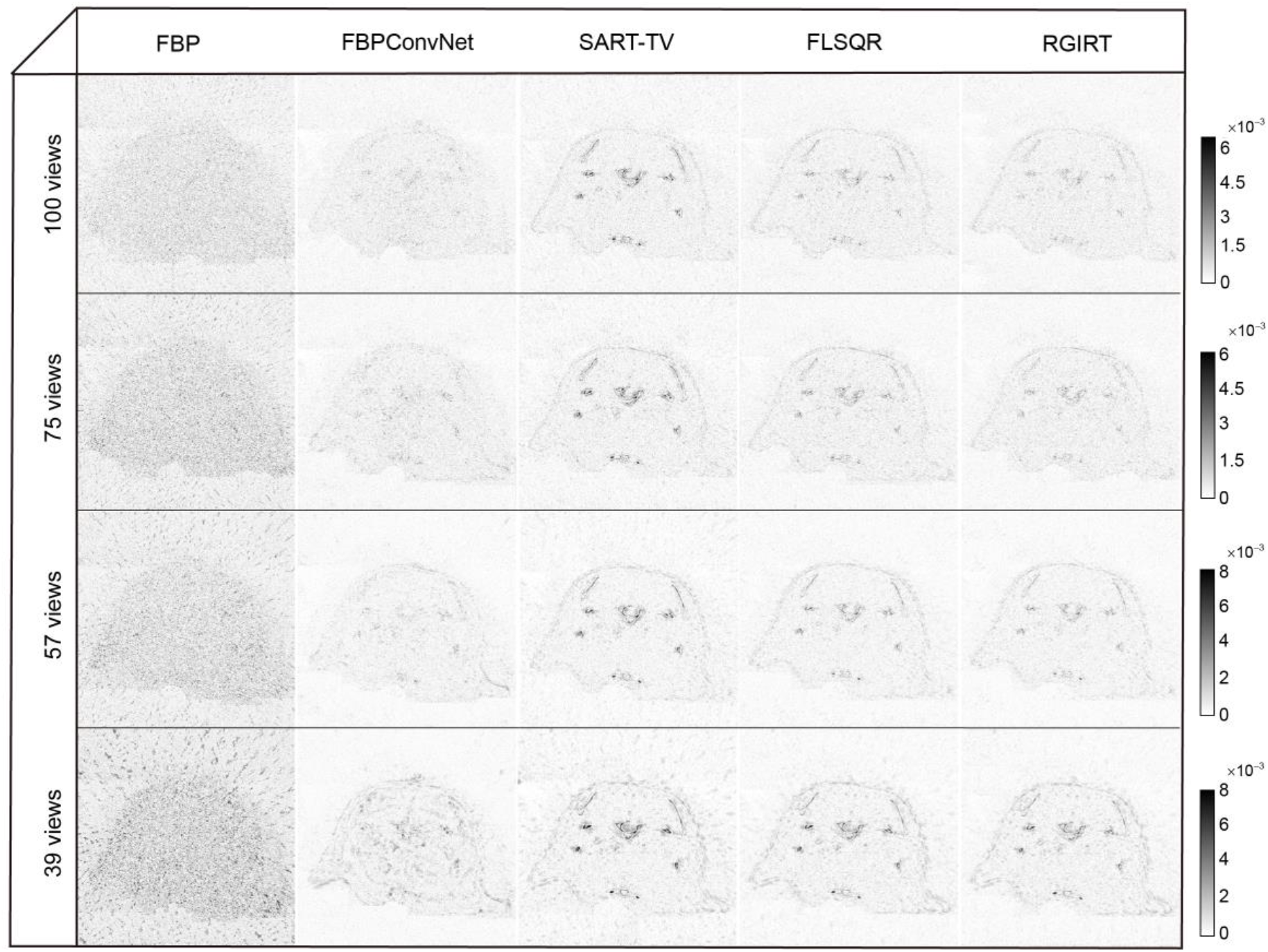
The difference images between the reference image and the sparse-view images in Fig. 2. Columns from left to right are difference images from FBP, FBPConvNet, SART-TV, FLSQR and RGIRT, respectively. Images from top to bottom show difference results for 100, 75, 57, and 39 projection views, respectively.

Fig. 5 shows the comparison of line intensity profiles of various methods passing through the yellow dash line (see Fig. 2) in the 57-view case. It is clear that the line intensity profile from RGIRT resembles most closely to the one from the reference CT image, especially for the peak and valley in the zoomed region. This demonstrates the advantage of our method over the other methods on persevering edge and small features in the images.

**Fig. 5.**
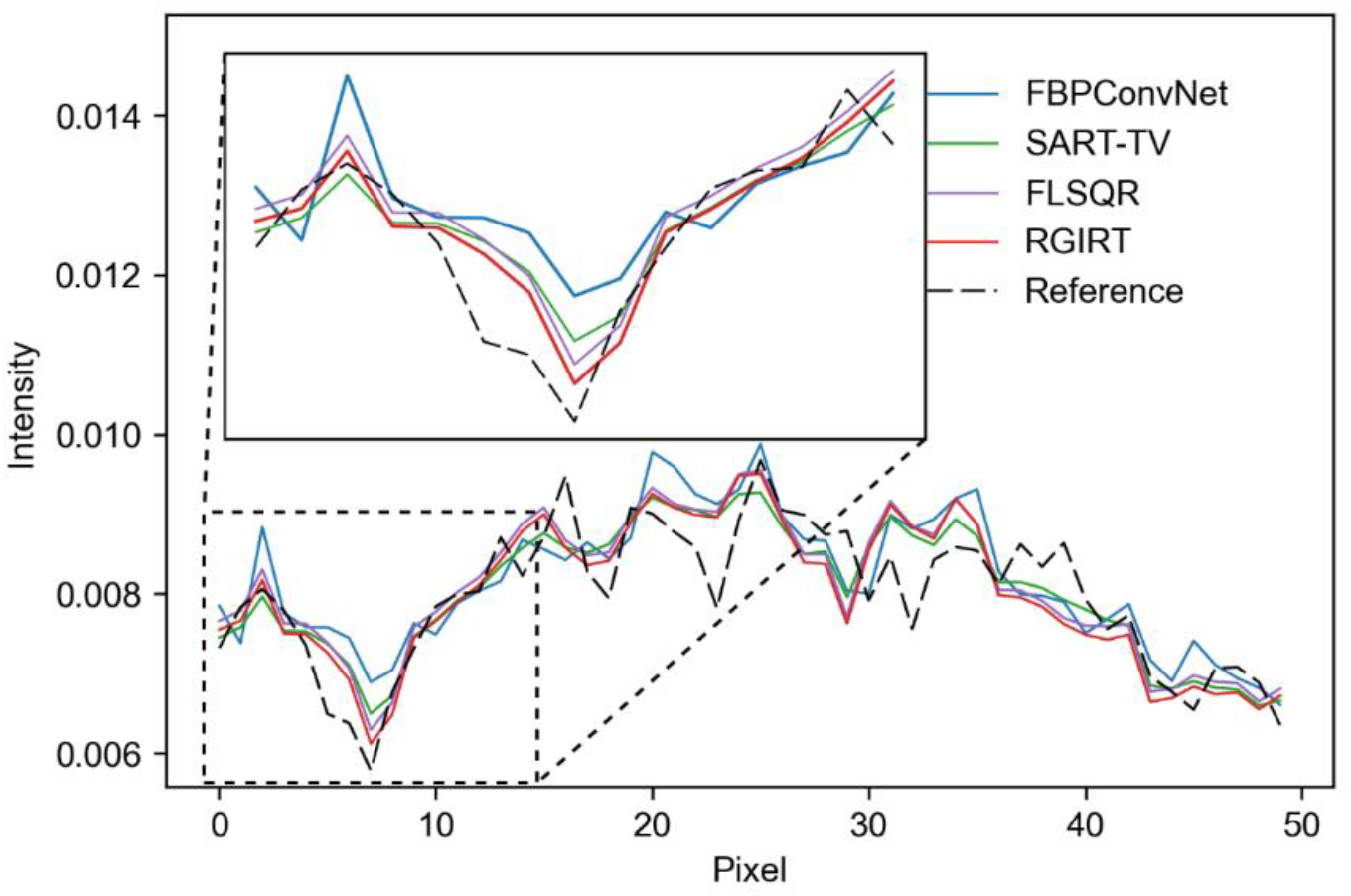
Line intensity profiles for the yellow dashed line in Fig. 2 (a3) (57 projection views). The inset shows the line from the our RGIRT method is closer to the reference line when compared to the other methods.

We also carried out quantitative analysis for the sparse-view cardiac micro-CT reconstructions and the results are presented in Table 1. As shown in Table 1, RGIRT and FBPConvNet clearly outperformed the other methods with smaller MSE and higher SSIM and PSNR. The FBPConvNet method has the best MSE among many sparse-view cases, mainly because it uses MSE as the loss function. However, as the aforementioned zoomed images (Fig. 3) clearly show, FBPConvNet has a poor performance in reducing the overall noise and noise textures, which led to the loss of small image details. Thus, although the FBPConvNet method provides good quantitative results, it cannot provide visual results as good as RGIRT. Compared to FBPConvNet our RGIRT method can consistently provide better visual results and quantitative results. Compared to the other reference methods, the superiority of RGIRT is more obvious when it comes to fewer projection views. Therefore, we can conclude that RGIRT provides the most accurate reconstruction results for sparse-view CT.

**Table 1.**
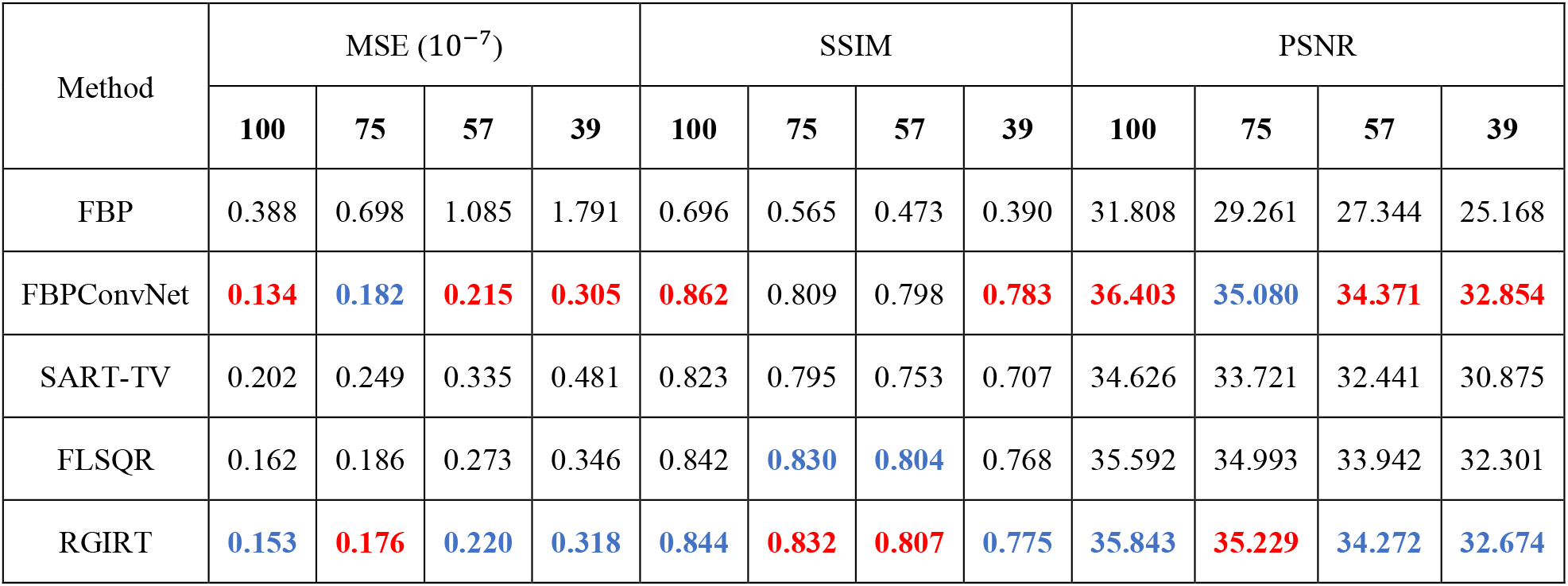
Quantitative results for sparse-view mouse cardiac micro-CT image. The best results are marked in red, and the second-best results are marked in blue.

| Method | MSE ($10^{-7}$) | | | | SSIM | | | | PSNR | | | |
| --- | --- | --- | --- | --- | --- | --- | --- | --- | --- | --- | --- | --- |
|  | 100 | 75 | 57 | 39 | 100 | 75 | 57 | 39 | 100 | 75 | 57 | 39 |
| FBP | 0.388 | 0.698 | 1.085 | 1.791 | 0.696 | 0.565 | 0.473 | 0.390 | 31.808 | 29.261 | 27.344 | 25.168 |
| FBPConvNet | **0.134** | **0.182** | **0.215** | **0.305** | **0.862** | 0.809 | 0.798 | **0.783** | **36.403** | **35.080** | **34.371** | **32.854** |
| SART-TV | 0.202 | 0.249 | 0.335 | 0.481 | 0.823 | 0.795 | 0.753 | 0.707 | 34.626 | 33.721 | 32.441 | 30.875 |
| FLSQR | 0.162 | 0.186 | 0.273 | 0.346 | 0.842 | **0.830** | **0.804** | 0.768 | 35.592 | 34.993 | 33.942 | 32.301 |
| RGIRT | **0.153** | **0.176** | **0.220** | **0.318** | **0.844** | **0.832** | **0.807** | **0.775** | **35.843** | **35.229** | **34.272** | **32.674** |

### 3.2 Convergence analysis

For the three iterative methods (SART-TV, FLSQR, RGIRT), we analyzed their convergence behaviors from the following three aspects: whether it converges, the speed of convergence, and image error. This convergence analysis can be examined by plotting the MSE loss curves during iterations, which are shown in Fig. 6. This figure demonstrates the capability of the three iterative methods in stabilizing the semi-convergent behavior of the ill-posed problem in the 57-view case.

**Fig. 6.**
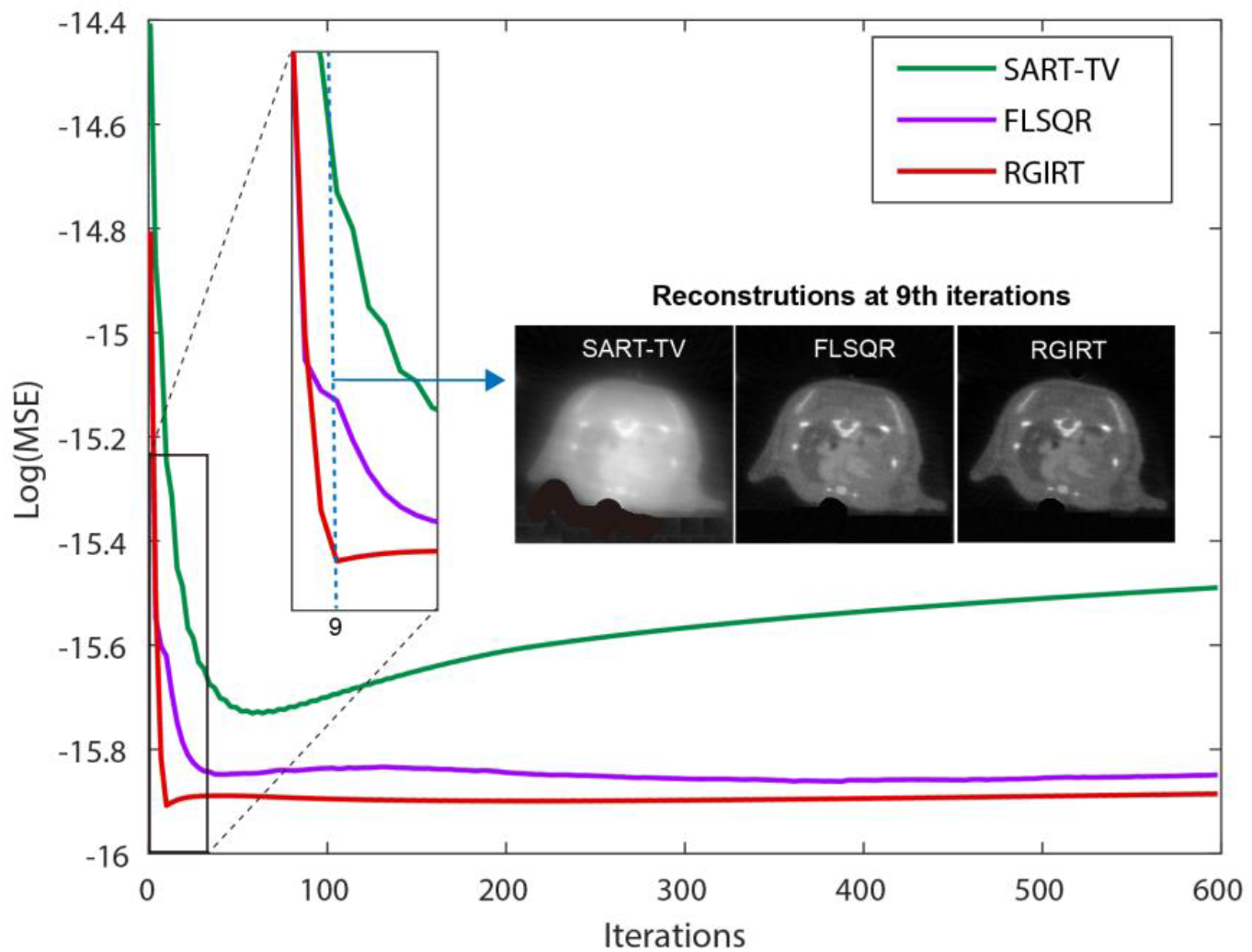
MSE loss curve of SART-TV (green), FLSQR (purple) and RGIRT (red) methods during iterations. The reconstruction results at the 9th iteration are visualized, showing that RGIRT converges faster than the others.

First, compared to FLSQR and RGIRT, SART-TV method has clear semi-convergent behavior, where MSE shows a trend of first decreasing and then slowly increasing as the number of iterations increases. As for the speed of convergence, it can be observed that RGIRT reaches the convergence point fastest with only 9 iterations. Furthermore, the image at the 9th iteration in Fig. 6 show that our RGIRT is able to obtain the most optimal result compared to the other methods. This is consistent with the performance of the MSE loss curve (see the zoomed area in Fig. 6). Combined with the fact that each iteration of RGIRT is to solve the dimensionality-reduced projected problem, therefore the computing time of RGIRT spent on one iteration is smallest among the three iterative methods. Thus, RGIRT is the fastest method to converge. In term of image error, RGIRT produced the lowest MSE in all the three iterative methods throughout the iterations. Therefore, among the three iterative methods studied in this work, RGIRT can provide the most accurate result with the fastest convergence speed.

### 3.3 Computational cost

Computational cost is an important factor for any reconstruction algorithm. All the algorithms in this work were implemented using MATLAB except the FBPConvNet, which was implemented with PYTHON. Considering that the training process for a DL method is very time consuming, the pre-training time was not included in comparison. Although the algorithms were realized using different programming languages, the efficiency can be roughly compared on a same computer. The MSE, SSIM and reconstruction time from the different methods when reconstructing the images for the 57-view case are shown in Fig. 7. All metrics in Fig. 7 were normalized to the largest value from various methods. For example, the SSIM of RGIRT is scaled to 1, and the SSIMs of other methods are normalized to the SSIM of RGIRT. The average reconstruction time for FBPConvNet, SART-TV, FLSQR, and RGIRT is 1.57, 84.66, 5.33, and 3.89 seconds per CT slice, respectively. All the reconstructions were conducted on the same PC workstation (Intel Xeon Gold 6248R CPU @ 3.00GHz, 128GB RAM and Nvidia Quadro RTX 4000 GPU card). Our RGIRT method clearly owns a considerable advantage in computational efficiency compared to the other iterative methods.

**Fig. 7.**
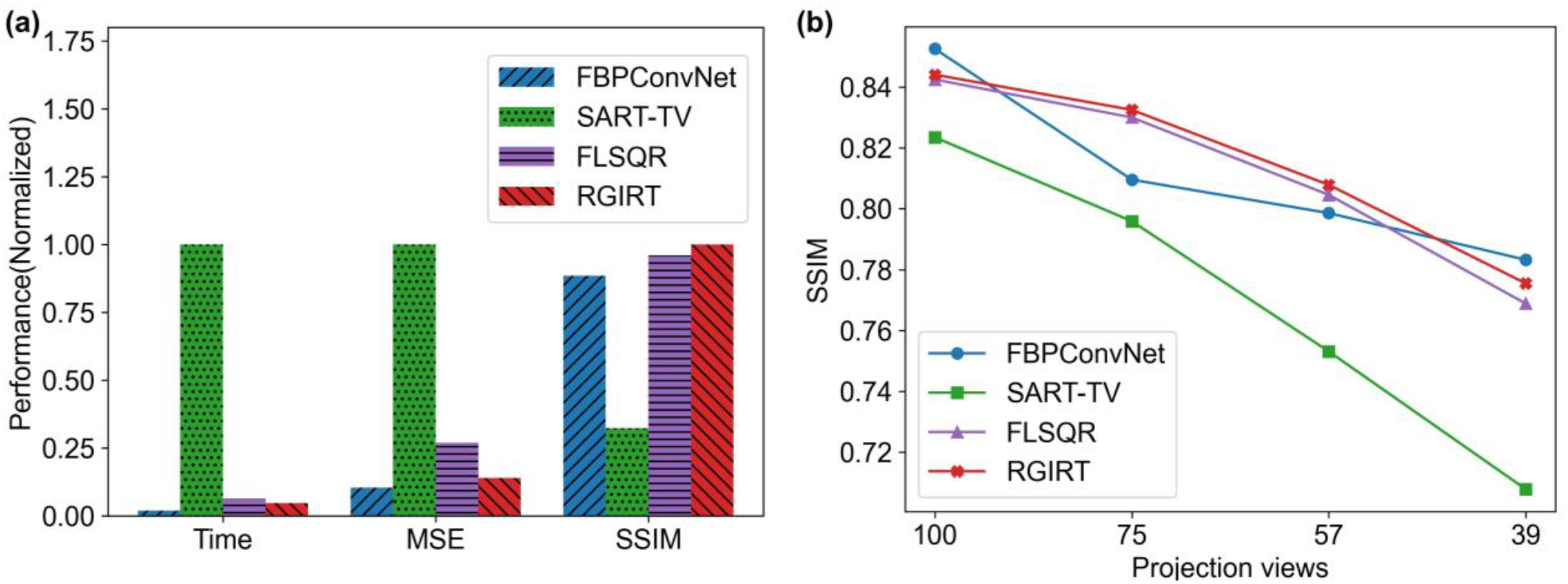
(a) Quantitative evaluation of different reconstruction methods in the 57-view case. (b) Variation of SSIM values with different projection views and reconstruction methods. RGIRT and FLSQR achieve a similarly smoother trend than FBPConvNet and SART-TV when the number of views changes.

## 4. Discussion and Conclusion

Although DL-based sparse-view CT reconstruction methods have gained increasing popularity in recent years, the interpretability and generalizability of trained networks remain unresolved issues in clinical applications [40]. A feasible strategy for resolving these issues is to combine deep learning networks with model-based iterative reconstruction methods [41]. The focus of this study is to develop a fast, accurate, and stable iterative reconstruction algorithm for sparse-view CT, which is beneficial for the future research into sparse-view CT reconstruction methods that leverage the synergistic powers of data-driven and model-driven methods. The proposed RGIRT algorithm is based on an inner-outer iteration framework, where FLSQR is implemented in the inner iteration and a restarted iterative scheme is applied in the outer iteration. Overall, RGIRT outperforms other reference methods (FBPConvNet, SART-TV, FLSQR) in terms of reconstruction accuracy and computational efficiency, while having a convergence guarantee demonstrated by experimental data and theoretical analysis.

Relative to other sparse-view CT reconstruction methods, the most important advantage of RGIRT is its convergence guarantee. Benefited from the unique design of inner-outer iteration framework, RGIRT employs a restarted iterative scheme in the outer loop to iteratively minimize the residual and refine the solution using the result obtained from the inner loop, thereby ensuring convergence. Classic iterative methods such as SART-TV only have semi-convergent behavior and do not have convergence guarantee (see Fig. 6). Similarly, it has been shown that during network training practical DL networks fail to converge to optimum solutions [42]. We believe the convergence guarantee of our RGIRT framework would be valuable in designing hybrid learning methods which will have the learning power of DL network and the convergence guarantee of RGIRT. This would certainly improve the explainability and generalizability (i.e., robustness) of hybrid DL methods.

Other two relative advantages of RGIRT are more accurate reconstruction results and higher convergence speed compared with reference methods. RGIRT can deliver more accurate reconstruction results, mainly because it enables efficient and automatic regularization parameter estimation via weighted generalized cross-validation (WGCV) [34] during the iterative process, thus underpinning a more reliable and precise outcome. As seen from the results in section 3.1, RGIRT is significantly better than the other methods, especially for the case with 57 projection views. The variation of SSIM with decreasing numbers of projection views indicates that RGIRT is distinguished by the lowest gradient of change, providing greater stability in more ill-posed cases with fewer projection views compared to other methods (see Fig. 7(b)). Furthermore, RGIRT features a faster convergence speed, mostly because it inherits the superior efficiency of the flexible Golub-Kahan bidiagonalization method, where the size of the projected problem is much smaller than the full-sized problem. The optimization procedure of RGIRT only requires matrix-vector multiplication in the iterative projection.

Our work is subject to following limitations. Firstly, RGIRT is currently designed for a certain type of regularization, i.e., *ℓ*_1_-norm regularization. In fact, other sparsity-encouraging prior knowledge can be adopted, such as *ℓ*_0_-norm regularized dictionary learning method [43]. It is obvious that more complex and advanced priors may improve the reconstructed image quality, although they are more computationally demanding. In practice, one would balance reconstruction performance and computational cost. Secondly, the inner-outer double iteration scheme of RGIRT could cause some computational complexity, especially when the convergence speed is slow for some special CT geometries. In this study we mainly focused on the short-scan in fan-beam geometry, since for a cone-beam CT scanner the short-scan is advantageous in terms of decreased mechanical manipulation, increased scanning flexibility, and shortened acquisition time.

In conclusion, using an inner-outer iteration framework with FLSQR implemented in the inner loop and restarted iterative scheme applied in the outer loop, RGIRT has consistently produced better results than representative deep learning and iterative reconstruction methods, including FBPConvNet, SART-TV, and FLSQR. The high-quality reconstruction of our method, together with its fast and stable convergence, renders it promising for sparse-view CT image reconstruction.

## Data Availability

All data produced in the present study are available upon reasonable request to the authors.

## Data Availability Statement

All data produced in the present study are available upon reasonable request to the authors.

## Acknowledgements

This work was partially supported by National Natural Science Foundation of China under grants 12101406 (JJ), 62105205 (WR), 62273238 (GC), Shanghai Science and Technology Innovation Program under grant 21YF1429100 (JJ), and start-up funding of ShanghaiTech University (WR, GC).

## Ethics declarations

### Competing interests

The authors declare no competing interests.

## Appendix A. Proof of convergence of the residual

Proof:

The residual at (*ℓ* + 1) −th outer iterations is

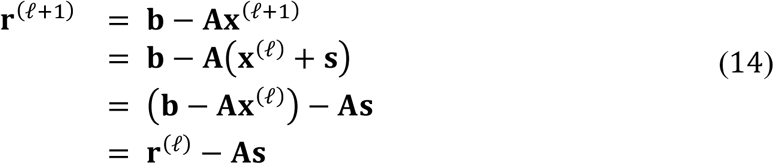

where **s** denotes the solution of the inner problem at *ℓ*-th outer iterations formulated as (4) in the main text and that **0** (zero vector) is in the solution space of **s**,

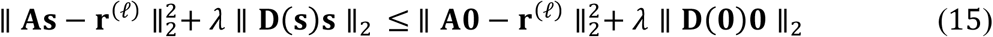

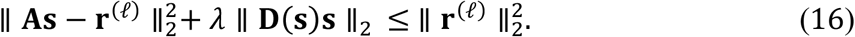

Meanwhile, due to *λ* > 0,

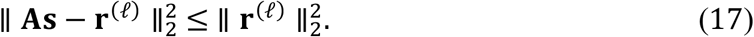

Combining the relationship of (14) and (17), we can obtain

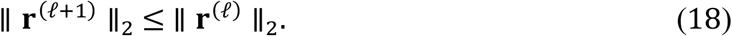

## Notes

### Competing Interest Statement

The authors have declared no competing interest.

